# Cortical GABAergic neuron dysregulation in schizophrenia is age dependent

**DOI:** 10.1101/2024.10.23.24315986

**Authors:** Daniel Kiss, Xiaolin Zhou, Keon Arbabi, Alex Gonzalez Segura, Nicole Endresz, Daniel Felsky, Andreea O. Diaconescu, Etienne Sibille, Shreejoy J Tripathy

## Abstract

Cortical GABAergic neuron dysregulation is implicated in schizophrenia (SCZ), but it remains unclear if these changes are due to altered cell proportions or per-cell mRNA changes. We analyzed bulk and cell type-specific RNAseq data from 1,302 individuals to assess SCZ- and age-associated changes in GABAergic interneurons across two neocortical regions. We found that younger SCZ cases (age < 70) showed reduced parvalbumin (PVALB) and somatostatin (SST) cell proportions, while older SCZ cases showed increased proportions compared to controls. Earlier onset SCZ, associated with more severe clinical symptoms, was linked to greater reductions in these cell types. Additionally, there was cohort-specific evidence for reduced per-cell PVALB and SST mRNA in SCZ. Our findings underscore the importance of age-stratified analyses in SCZ, suggesting that distinct pathological processes underlie GABAergic neuron dysregulation across different age- and symptom-severity groups and warranting tailored therapeutic approaches.

## Introduction

Schizophrenia (SCZ) is a debilitating neuropsychiatric condition affecting approximately 1% of North Americans ^1^. SCZ is characterized by positive symptoms (e.g., delusions, hallucinations), negative symptoms (e.g., avolition, asociality), and cognitive impairments ^1–4^. The SCZ phenotype is heterogeneous, with factors such as age of onset, sex, genetics and lifestyle, impacting severity and prognosis ^5,6^. On average, in North America, SCZ is associated with a decreased life expectancy, with an earlier diagnosis leading to more severe symptoms and cognitive impairments ^7–10^.

Studies have explored the associations between SCZ and cell type-specific abnormalities in the brain, particularly in cortical GABAergic signaling networks ^11–13^. Changes in the functioning of GABAergic interneurons have been linked to working memory and cognitive impairments in SCZ ^11^, contributing to a wide range of symptoms through imbalances in excitation and inhibition. Alterations in two particular GABAergic interneuron subtypes—parvalbumin (PVALB) and somatostatin (SST) expressing interneurons—are most strongly associated with SCZ ^11,14,15^. Fast-spiking PVALB interneurons regulate the activity of pyramidal neurons by providing inhibition and controlling the precise timing of neural firing within cortical circuits ^16,17^. They are implicated in the maintenance of gamma oscillations in the dorsolateral prefrontal cortex (DLPFC) and are closely tied to cognitive processes, including working memory and cognitive flexibility ^18^. The DLPFC and cingulate cortex have been specifically implicated in SCZ pathology, with fMRI studies showing abnormal activations in these areas thought to underlie cognitive impairments in SCZ ^19^. SST interneurons provide dendritic inhibition to pyramidal neurons and modulate theta network oscillations ^18,20^. Deficits in SST interneurons in mammals are linked to dysregulated attention, episodic memory, and spatial navigation ^11,14,20,21^, and have been observed across multiple brain disorders as well as in healthy aging ^22–24^, suggesting SST cell-intrinsic vulnerability ^25^.

Despite evidence implicating interneuron alterations in SCZ, it remains unclear whether these associations are the result of cell type-specific molecular (e.g. mRNA) alterations or changes in cellular abundance and density. For example, a meta-analysis by Kaar et al. (2019) supports reduced PVALB cell density in SCZ and suggests that PVALB mRNA levels within PVALB cells are not significantly changed ^26^. Similarly, Toker et al. (2019) found decreases in PVALB cell proportions in SCZ through a large transcriptomic meta-analysis primarily employing bulk tissue deconvolution ^27^. More recently, Batiuk et al. (2022) utilized single-nucleus RNAseq (snRNAseq) to reveal a decreased abundance of specific interneuron subtypes in SCZ, particularly upper-layer SST and PVALB cells ^28^. In contrast to observations of altered cell type densities in SCZ, a recent study by Dienel at al. (2023) used in situ hybridization (ISH) techniques to quantify the density of PVALB and SST expressing cells as well as the endogenous mRNA expression of PVALB and SST mRNA levels in these cells ^29^. Altered interneuron cell densities were not observed, whereas PVALB and SST mRNA per cell were found to be reduced in SCZ. These findings are consistent with other studies which have found evidence for reduced PVALB mRNA expression per cell but unchanged cellular densities in SCZ ^30–32^.

Sabunciyan (2019) identified interactions between age and diagnosis in SCZ at the transcriptomic level, showing that gene expression differences between SCZ and healthy controls vary with age ^33^. Specifically, this study suggests that the most considerable SCZ-associated differential expression occurs in mid-life (ages 40-60 years), diminishes in late life (ages >60), and may contribute to “premature brain aging” in SCZ ^34,35^. Similarly, recent studies, including our own, have linked healthy aging to reductions in SST and PVALB interneurons ^22^. Given the parallels between SCZ and aging, particularly in cell type proportion changes, it is crucial to investigate age-disease interactions in SCZ. Notably, the association between SCZ-related PV and SST interneuron pathology and factors like age of onset, age at death, and disease severity remains unexplored.

In this study, we aim to clarify the contributions of cell type-specific molecular alterations and changes in cellular proportions to the pathophysiology of schizophrenia, with a particular focus on PVALB and SST interneurons. We hypothesize that the dysregulation of these interneurons in schizophrenia is age-dependent, with distinct patterns of cell proportion and mRNA expression changes across different age groups. Additionally, we explore the relationship between these alterations and clinical factors such as age of SCZ onset and age at death, positing that early-onset schizophrenia may be associated with more pronounced interneuron deficits. By integrating data from bulk and cell type-specific RNA sequencing, we seek to provide a more nuanced understanding of the molecular and cellular mechanisms underlying schizophrenia and how they interact with the aging process.

## Methods

### Transcriptomic Datasets

We utilized twelve post-mortem transcriptomics datasets, reflecting all of the post-mortem datasets reflecting adult schizophrenia cases and controls in the PsychENCODE and CommonMind consortia ^36–41^ (detailed in Table 1). Together, these datasets reflect data from six institutional cohorts and/or brain banks (Pitt, MSSM, Penn, NIMH_HBCC / LIBD, SMRI, and McLean); two sampled brain regions (dorsolateral prefrontal cortex, DLPFC, and anterior cingulate cortex, ACC); and three genomics sampling techniques (bulk RNAseq, single-nucleus RNAseq, and fluorescence in situ hybridization followed by LCM-seq). To ensure that samples were reflective of the adult human neocortex from SCZ cases and controls, we filtered datasets to those spanning 15 to 90+ years of age at death and excluded data from donors without a control or SCZ case diagnosis.

**Table 1.** Summary of RNAseq datasets. Dataset / Cohort Name denotes name of dataset used in this study. Brain area denotes neocortical area sampled in the dataset. Cohort / Brain Bank denotes the name of the cohort and brain bank: MSSM (Mount Sinai School of Medicine), Penn (University of Pennsylvania), Pitt (University of Pittsburgh), HBCC / LIBD (National Institutes of Mental Health Human Brain Collection Core / Lieber Institute for Brain Development), SMRI (Stanley Medical Research Institute), McLean (McLean Hospital). RNA Type denotes type of RNAseq data collected, either Bulk tissue RNAseq, single-nucleus RNAseq (snRNAseq), or laser capture microdissection-RNAseq (LCMseq). Synapse ID denotes identifier at synapse.org where data were obtained.

| Dataset / Cohort Name | Brain Area | Cohort / Brain Bank | % Male | Age (SD) | RNA Type | # Case | # Control | Synapse ID |
| --- | --- | --- | --- | --- | --- | --- | --- | --- |
| MSSM (bulk) | DLPFC | MSSM | 59.6 | 72.8 (14.6) | Bulk | 149 | 163 | syn22344686 |
| Penn | DLPFC | Penn | 43.2 | 74.2 (13.9) | Bulk | 58 | 37 | syn22344686 |
| Pitt | DLPFC | Pitt | 79.2 | 48.7 (13.9) | Bulk | 57 | 49 | syn22344686 |
| NIMH_HBCC | DLPFC | HBCC / LIBD | 71.3 | 43.9 (16.6) | Bulk | 96 | 186 | syn22344107 |
| LIBD_szControl | DLPFC | HBCC / LIBD | 67.1 | 44.9 (16.2) | Bulk | 175 | 223 | syn22344107 |
| GVEX | DLPFC | SMRI | 75.1 | 44.5 (10.3) | Bulk | 94 | 75 | syn4590909 |
| Pitt_ACC | ACC | Pitt | 71.8 | 47.9 (13.8) | Bulk | 58 | 78 | syn22344686 |
| MSSM_ACC | ACC | MSSM | 58.6 | 73.1 (14.6) | Bulk | 130 | 143 | syn22344686 |
| Penn_ACC | ACC | Penn | 45.5 | 73.8 (13.8) | Bulk | 43 | 23 | syn22344686 |
| McLean | DLPFC | McLean | 66.3 | 63.2 (16.9) | snRNAseq | 24 | 24 | syn25946131 |
| MSSM (snRNA) | DLPFC | MSSM | 50.0 | 72.7 (16.5) | snRNAseq | 41 | 51 | syn25946131 |
| Pitt_sgACC<br>FISH+LCMseq | sgACC | Pitt | 50.0 | 48.3 (9.8) | LCM-seq | 19 | 19 | NA |

We note that some tissue samples in different studies were occasionally collected from the same donor, albeit from different brain regions or with different techniques (e.g., bulk and snRNAseq collected from the same donor as part of different studies). Table 1 shows the source cohort / brain banks for each included dataset.

The Genetic Variants Affect Brain Gene Expression (BrainGVEX) study provided bulk RNAseq data from the DLPFC of 169 donors (94 cases and 75 controls with a mean age of 44.5 years and a standard deviation of 10.3). Though the study comprises multiple cohorts from the Stanley Brain Collection (SMRI Array, SMRI Consortium, SMRI New, SMRI Extra), due to the smaller sample sizes in each cohort, we opted to combine them into a single large cohort, herein referred to as GVEX.

We analyzed cell density and cell type-specific expression data from 38 donors (19 cases and 19 healthy controls) obtained using fluorescence in situ hybridization followed by laser-capture microdissection RNA sequencing (FISH+LCMseq) ^39^ collected from a subset of age and sex-matched donors from the Pitt cohort.

#### Tissue Collection and RNA Sequencing Protocols

Tissue preparation and RNA sequencing protocols differed between studies and cohorts. Information regarding the management and processing of post-mortem DLPFC and ACC RNAseq samples have been previously published for each dataset ^36–41^.

#### Bulk RNAseq Pre-Processing and Quality Control

Bulk RNAseq counts for each sample were downloaded from publicly-available sources and aggregated into count matrices. Matrices were converted into counts per million using the cpm function from edgeR ^42^ and log2 transformed with a prior count of 0.1. Genes with low standard deviation (SD < 0.1) were filtered from subsequent analyses. No additional outliers were identified at the gene expression level, and thus no samples were excluded.

### Reference cell type taxonomy

The reference neocortical cell type taxonomy used for this study was obtained from the Allen Institute for Brain Sciences (AIBS) “Multiple Cortical Areas - Smart-seq (2019)” dataset in August 2020 through the website https://portal.brain-map.org/atlases-and-data/rnaseq/human-multiple-cortical-areas-smart-seq. The sample collection and data analysis methods are described in ^43,44^. This dataset contains 49,495 nuclei that were sampled from several regions of the human neocortex, including the medial temporal gyrus, anterior cingulate cortex, primary visual cortex, primary motor cortex, primary somatosensory cortex, and primary auditory cortex. The data was obtained using ultra-high depth single-nucleus RNA sequencing. We applied cell type taxonomies at the “subclass” level via the “subclass_label” field, providing us with an intermediate level of detail that splits broad excitatory and inhibitory cell classes while preserving moderate cell type granularity. In total, this dataset classifies neocortical cells into 19 distinct cell types, encompassing all major expected cell types in the neocortex.

### Estimation of Bulk RNAseq-derived Relative Cell Type Proportions (rCTPs)

For bulk RNAseq datasets, we performed relative cell type proportion estimation using mgpEstimate from the MarkerGeneProfile (MGP) R package as described by ^27,45^. We made use of our prior list of cell type-specific marker genes for each cell type as described previously ^46^. MGP estimates were interpreted as relative cell proportion estimates (rCTPs), which serve as a proxy for cell type abundance in each sample. rCTPs were z-score normalized within each dataset for all subsequent analyses.

We also estimated cell type proportions using dtangle, a well-established deconvolution method for estimating cell type proportions from bulk-RNAseq data ^47,48^, as a second deconvolution algorithm. dtangle differs from the MGP method described above in that it estimates absolute cell type proportions, not relative proportions. Specifically, we used the find_markers() function with the “ratio” argument provided by dtangle to find marker genes for each cell type. We included the top 10% of highly expressed genes as markers per cell type. The AIBS single-nucleus RNA-seq dataset ^43^ was used as our reference dataset for cell type gene-expression profiles for dtangle.

### Case Control Single-nucleus and LCM-based RNAseq

#### Single-nucleus RNAseq Pre-processing and Quality Control

Single-nucleus datasets from the Ruzicka study ^40^ for each sample were downloaded from Synapse (<u>syn25946131</u>) and aggregated into count matrices for each of the two cohorts used in this study, McLean and MSSM. Samples were removed if less than 500 nuclei passed quality control criteria.

Due to differences in cell type taxonomies between this single nucleus dataset and those of the other analyses in this paper based on ^43^, we mapped the cell type definitions of each nucleus to that of our reference cell type taxonomy described above. Reference-based label transfer was performed using Seurat ^49^ to project reference data onto the query object. Anchors were identified between the datasets, and the reference PCA structure was projected onto the query. The TransferData() function was then used to classify query cells based on reference cell type labels, generating predicted cell identities and scores, which were added to the query data (Satija Lab, n.d.). Donors were discarded from downstream analyses if less than 500 total quality-control passing nuclei were sampled.

#### Single-Nucleus RNAseq-derived Cell Type Proportions

For single-nucleus RNAseq datasets, we quantified cell type proportions from snRNAseq datasets (snCTPs) using the same approach to our prior analyses ^22,46^. Specifically, we quantified the total count of nuclei annotated to each cell type perdonor, and then normalized it by the total number of nuclei sampled from that donor (i.e., snCTPs reflect absolute proportions of each cell type per donor).

#### Generation of cell type-specific pseudobulk expression profiles from single-nucleus RNAseq datasets

Following cell type mapping, to provide a more robust representation of gene expression at the cell type level for each donor, the label-transferred data was pseudo-bulked using Seurat’s AggregateExpression() function ^49^, which calculates the aggregated gene expression profile for each cell type by grouping cells based on their assigned cell type labels. A benefit to pseudo-bulking is that it helps mitigate the sparsity and noise inherent in single-cell data, providing a more robust representation of gene expression at the cell type level ^50^.

#### Comparison of bulk deconvolution methods with cell proportions from single-nucleus RNAseq and microscopy-based cell densities

Multiple methods were used in this study for assessing the accuracy of cell type deconvolution algorithms. We first identified brain datasets where bulk tissue RNAseq and a second data modality, e.g. snRNA-seq, were collected from the same brain region from multiple donors.

We identified two such scenarios: first, we noticed that bulk RNAseq from the DLPFC from a subset of donors in the MSSM cohort as part of the CommonMind Consortium samples originated from donors with snRNAseq data collected from the prefrontal cortex (Brodmann Areas 9, 10, or 46) as part of the Ruzicka study ^40^.

Second, we observed a small set of overlapping donors from the Pitt cohort of the CommonMind Consortium samples with bulk RNAseq sampled from the ACC that also had FISH microscopy-based cell densities and LCM-seq collected from the subgenual ACC as part of our prior Arbabi et al study ^39^. In making these comparisons, we also made use of data from 19 donors with bipolar disorder who were commonly sampled between these two studies. We note that samples with cell densities greater than three times the interquartile range were excluded to eliminate outliers prior to making comparisons.

### Statistical Analyses

#### Association of Estimated Cell Type Proportions (CTP) with Schizophrenia Case/Control Status

To estimate associations of SCZ case/control status and other covariates with bulk and single-nucleus based CTP values, we used a number of statistical models.

First, for rCTPs estimated from bulk tissue samples from both DLPFC and ACC, we fit a series of linear models for each cell type and bulk RNAseq dataset. RNA integrity number (RIN), postmortem interval (PMI), and age at death were included as covariates and z-score normalized prior to analysis.

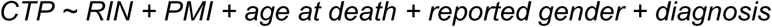

For each model, adjusted p-values (*p_adj_*) were obtained using False Discovery Rate (FDR) correction ^51^. For each dataset, CTP values were residualized for covariates and compared between SCZ and healthy controls using Mann-Whitney U tests ^52^.

#### Modeling Interactions Between Schizophrenia Diagnosis and Age at Death

We performed age-stratified mega-analyses (incorporating all bulk tissue datasets available) by associating CTPs with SCZ case/control status within each decade-at-death age bin per cell type. Since the youngest case of SCZ across all datasets had an age at death of 17, our first age bin included all subjects between the ages of 15 and 29. In aggregate, we performed a set of linear models stratifying for each cell type and each age bin by decade (ageDeath = 15-29 to ageDeath = 90+). This analysis was repeated by binning age into two categories, above and below donor age of death of 70 years.

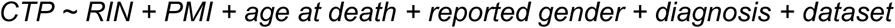

Additionally, we combined donors of all ages and performed a set of mega-analyses associating rCTPs with SCZ case/control status in each cell type, including interaction terms for ageDeath x diagnosis.

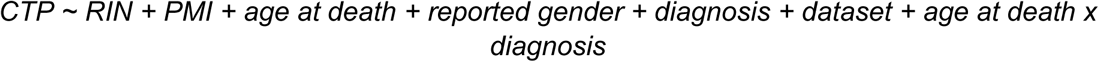

#### Analysis of Single Nucleus RNAseq snCTP Associations

For snCTP association with SCZ case/control status, we performed similar age-stratified linear models as for rCTPs using the same covariates, with RNA integrity (RIN) omitted due its lack of availability in all datasets. Additionally, due to smaller sample sizes, binning donors by decade of age at death was not feasible. We thus divided samples into two groups, above and below ageDeath = 70.

#### Analysis of Cell Type-Specific Densities based on Fluorescent In Situ Hybridization

We obtained FISH-based cell density information (cells/mm^2^) from Arbabi et al ^39^ and associated these cellular densities with SCZ case/control status. Since the oldest donor in our microscopy dataset had ageDeath = 68, we elected to omit any age-stratified analyses.

#### Association of Schizophrenia rCTPs with Age of Onset and Other Covariates

To assess cell type associations with age of onset and other covariates within SCZ cases, we first residualized rCTPs for RIN, PMI, age at death, and gender using the model below using SCZ cases only:

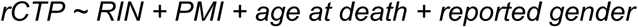

Residualized rCTPs were then correlated with SCZ age of onset and assessed by calculating Pearson’s coefficient. The association of SCZ rCTPs with age of onset was only performed in GVEX and LIBD_szControl cohorts, as age of onset information was not provided for the other datasets.

#### Association of Cell Type Specific Expression and Association with Schizophrenia Case/Control Status

Cell type-specific SCZ-related differential expression (DE) was quantified for snRNAseq by pseudobulking gene expression data per individual and cell type, according to cell type definitions used by Consens et al ^46^. Given our focus on GABAergic interneuron subtypes which tend to be somewhat rare, we excluded snRNAseq data for samples with fewer than 1000 total QC-passing nuclei. We performed standard differential expression analyses for each cell type for groups above and below ageDeath = 70 with limma-voom edgeR ^53,54^ using the following model:

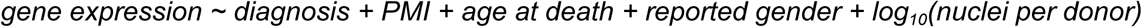

Models included SCZ disease state as its main predictor, and covaried for PMI, subject age at death, subject gender, cohort, and the base 10 logarithm of the number of QC-passing nuclei.

We also utilized differential expression summary statistics from our prior FISH+LCM-seq dataset from the sgACC brain region from the Pitt cohort from ^39^.

#### Multiple Comparisons Consideration

For each model, adjusted p-values were obtained using False Discovery Rate (FDR) correction ^51^ to account for comparisons across multiple cell types and also multiple genes.

## Results

### Proportions of GABAergic interneuron subtypes can be deconvolved from human brain bulk RNAseq data

Given the abundance of brain bulk RNAseq datasets from post-mortem SCZ cases and controls, we first assessed the accuracy of cell type deconvolution methods for estimating GABAergic interneuron subtype proportions. We compared deconvolution results using the Marker Gene Profile (MGP) method ^27,45^ and dtangle ^47^ with “gold standard” cell type proportions from single-nucleus RNAseq (snRNAseq) and microscopy-based cell densities.

Using data from the MSSM cohort which included both bulk RNAseq and snRNAseq from the DLPFC of the same donors ^40^, we observed that MGP-based relative cell type proportions (rCTPs) were correlated with single-nucleus proportions (snCTPs) for PVALB, SST, and VIP interneurons, with Pearson’s R values of 0.37, 0.61, and 0.41, respectively (Fig. 1B). Moreover, across all cell types, we found that MGP-based rCTPs were usually positively correlated with snCTPs (Fig. 1C). We found qualitatively similar findings using dtangle, an alternative method for cell type deconvolution (Supplementary Figure S1). Given these results, we opted to primarily utilize the MGP algorithm here, given our recent usage of this cell type deconvolution tool to understand changes in human Alzheimer’s disease and healthy aging ^34,46^.

**Figure 1.**
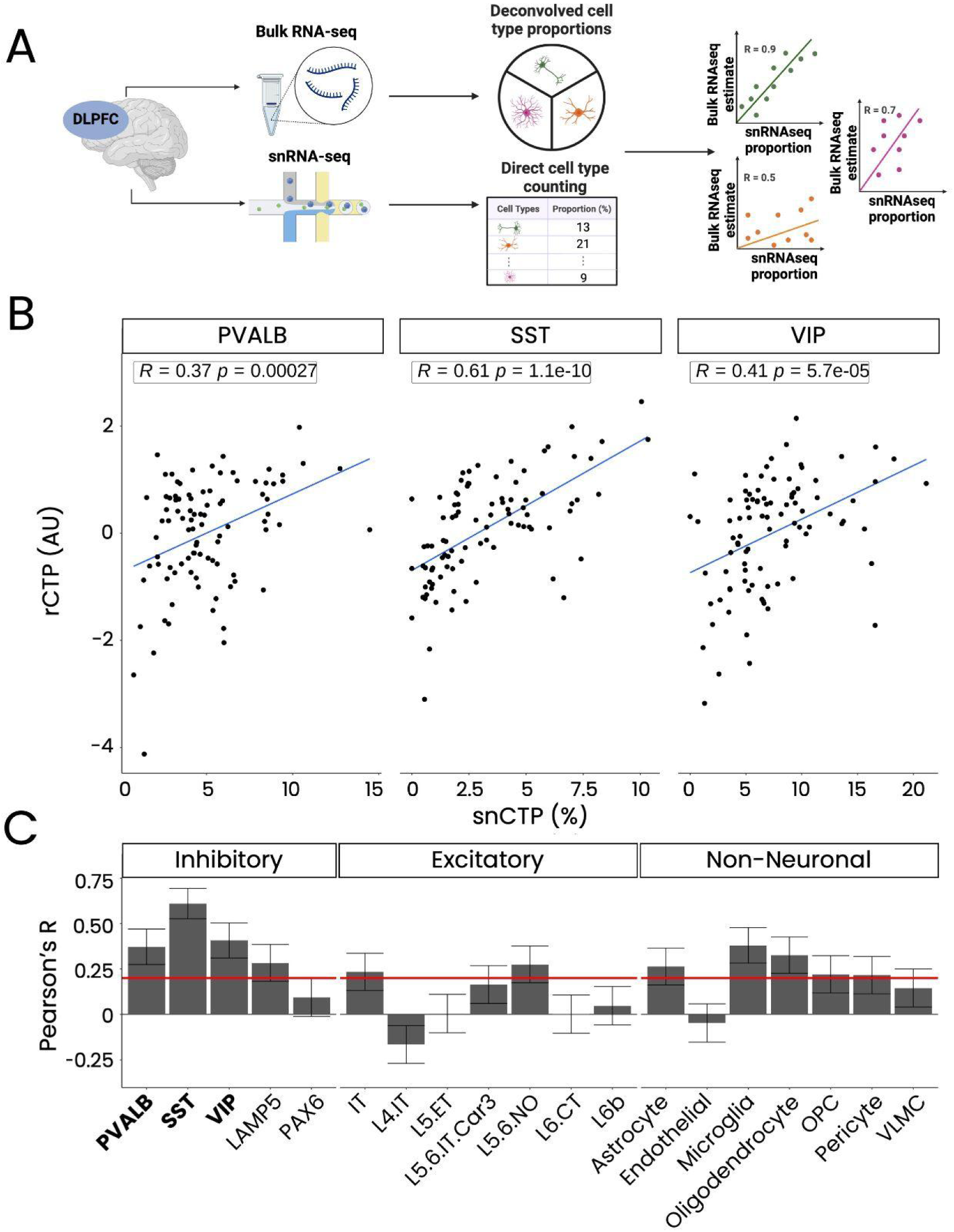
Concordance between cell proportion estimates from bulk tissue deconvolution and single-nucleus RNA-seq from matched tissue samples. (A) Schematic representation of bulk RNA-seq collection and cell proportion estimation via deconvolution (top), alongside single-nucleus RNA-seq (snRNA-seq) with direct cell proportion calculation through cell counting (bottom) for DLPFC samples from matched donors. The rightmost panel shows the calculation of concordance between bulk deconvolution and snRNA-seq proportions using correlation coefficients. Each dot represents a distinct individual, and colors indicate different cell types. (B) Scatterplots showing the correlation between single-nucleus-based cell type proportions (snCTPs; x-axis, % of total cells in the sample) and relative cell type proportions (rCTPs; y-axis, arbitrary units, AU) derived from bulk tissue deconvolution. Inset values show Pearson’s correlation coefficient (R) and corresponding p-values. (C) Bar plot displaying Pearson’s R values for various cell types (x-axis). Red line indicates Pearson’s R of 0.2 and error bars represent the standard error of the mean (SEM).

We further validated bulk tissue deconvolution against microscopy-based cell densities using the Pitt cohort ^39^, where we previously used stereological methods to quantify PVALB, SST, and VIP cell densities in the subgenual ACC via fluorescence in situ hybridization (FISH). Comparing these with rCTPs derived from bulk RNAseq data from the same donors, we found Pearson’s R values of 0.34, 0.40, and 0.38 for PVALB, SST, and VIP, respectively (Supplementary Figure S2).

Overall, our analyses suggest that bulk tissue deconvolution using RNAseq data from the human neocortex is a reliable method for estimating GABAergic interneuron subtype proportions, allowing for the assessment of inter-individual differences in these cell populations.

### Cohort-dependent differences in PVALB and SST cell proportions in SCZ

To assess whether SCZ is associated with differences in GABAergic interneuron subtype proportions, we analyzed nine datasets from five post-mortem cohorts within the CommonMind and PsychEncode consortia (Table 1). Bulk RNAseq data were collected from the dorsolateral prefrontal cortex (DLPFC) or anterior cingulate cortex (ACC). We used the MGP algorithm to estimate relative cell type proportions (rCTPs) for each major cell type in each dataset.

We first examined how rCTPs differed between SCZ cases and controls, adjusting for demographic and technical factors (Figure 2A). SCZ was associated with reduced PVALB and SST rCTPs in four of six DLPFC datasets (GVEX, NIMH, Pitt, and LIBD) at FDR < 0.10. In the Penn cohort, PVALB and SST rCTPs did not differ significantly between SCZ and controls, while the MSSM cohort showed increased PVALB rCTPs in SCZ but no change in SST rCTPs. VIP cells generally showed no significant differences, except for decreased rCTPs in the NIMH dataset (β = -0.40, FDR = .053). Similar patterns were observed in the ACC datasets, with SCZ-associated reductions in PVALB, SST, and VIP cells in the Pitt ACC dataset, and no significant changes in the MSSM and Penn ACC datasets (Supplementary Figure S3).

**Figure 2.**
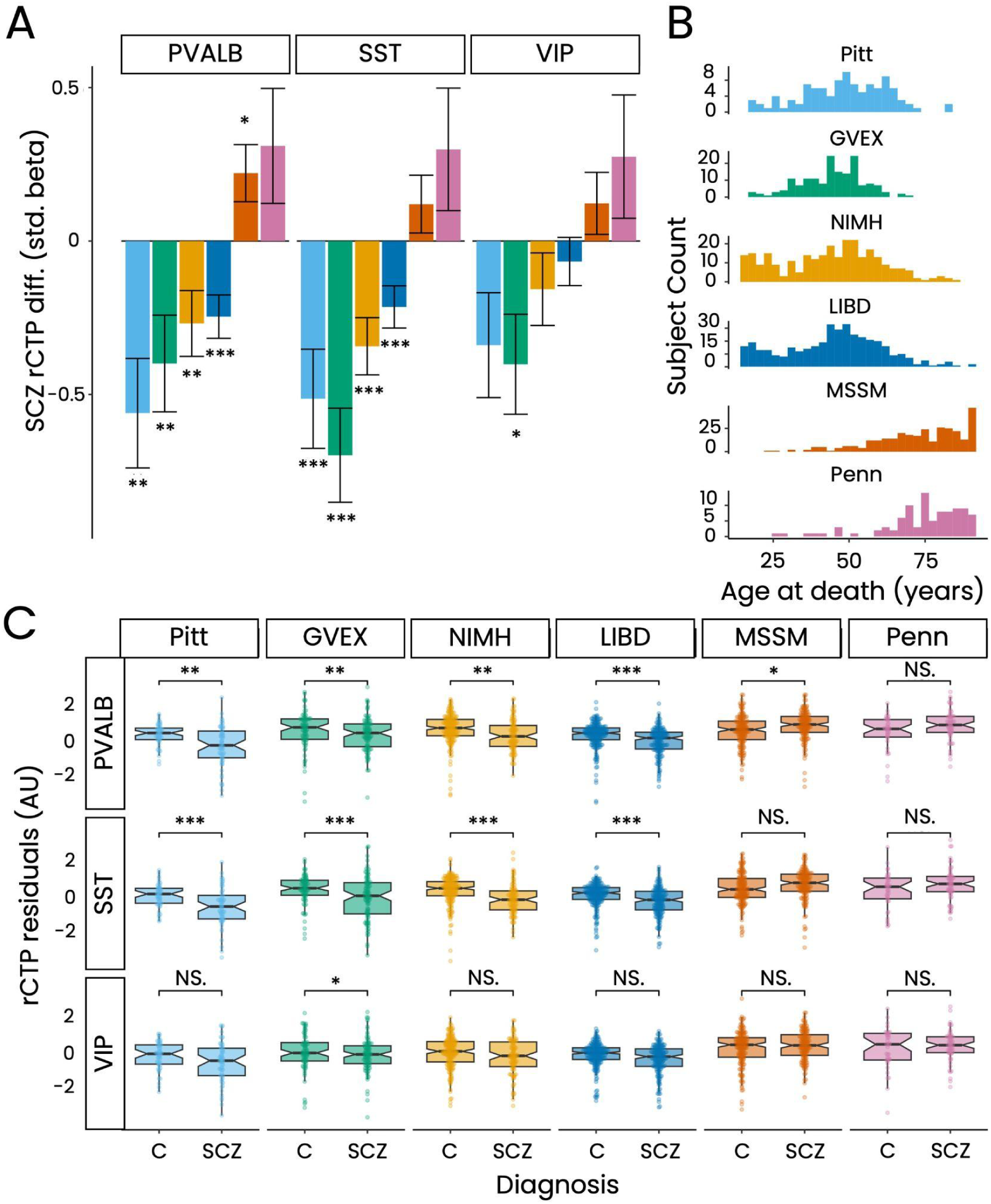
Differences in DLPFC interneuron proportions between schizophrenia and controls. (A) Bar plots showing standardized beta coefficients (β) for differences in relative cell type proportions (rCTPs) of PVALB, SST, and VIP interneurons between schizophrenia (SCZ) and control (C) groups across six bulk RNA-seq datasets. Positive values indicate increased proportions in SCZ, while negative values indicate decreases. Error bars represent standard error, and asterisks indicate significance based on false discovery rate FDR (* = 0.1, ** = 0.05, *** = 0.01). (B) Age at death distribution for each dataset. Datasets are ordered by increasing mean age in years. (C) Box plots of residualized rCTPs, after controlling for demographic and technical covariates, for PVALB, SST, and VIP interneurons across datasets. Asterisks reflect significance from panel (A), with “NS” for non-significant comparisons.

Upon investigating factors that might explain these differing results, we found that the cohorts varied widely in average participant age of death(Figure 2B). Datasets with younger donors; GVEX (mean age at death = 44.5), NIMH (mean age at death = 43.9), Pitt (mean age at death = 48.7), and LIBD (mean age at death = 44.9), tended to show SCZ-associated reductions in PVALB and SST rCTPs, whereas those with older donors in MSSM (mean age at death = 72.8) and Penn (mean age at death = 74.2) showed unchanged or increased rCTPs (Supplementary Figure S4). These results suggest that age is a significant factor in whether PVALB and SST rCTPs are observed as increased or decreased in SCZ.

### SCZ-associated changes in PVALB and SST cell proportions vary with individual age

Given our observation that SCZ-related changes in PVALB and SST rCTPs might be influenced by age at death, we directly assessed these effects across the aging spectrum. We combined rCTPs across all bulk RNAseq DLPFC datasets (Figure 3A) and ACC samples (Supplementary Fig. S5), adjusting for dataset differences. In controls, we observed a cross-sectional decrease in PVALB, SST, and VIP cell proportions with age, consistent with previous findings ^22,23^. However, in SCZ cases, rCTPs were more uniform and lower across donors and ages, suggesting a trend towards “accelerated” or “anticipated” aging in SCZ ^35^.

**Figure 3.**
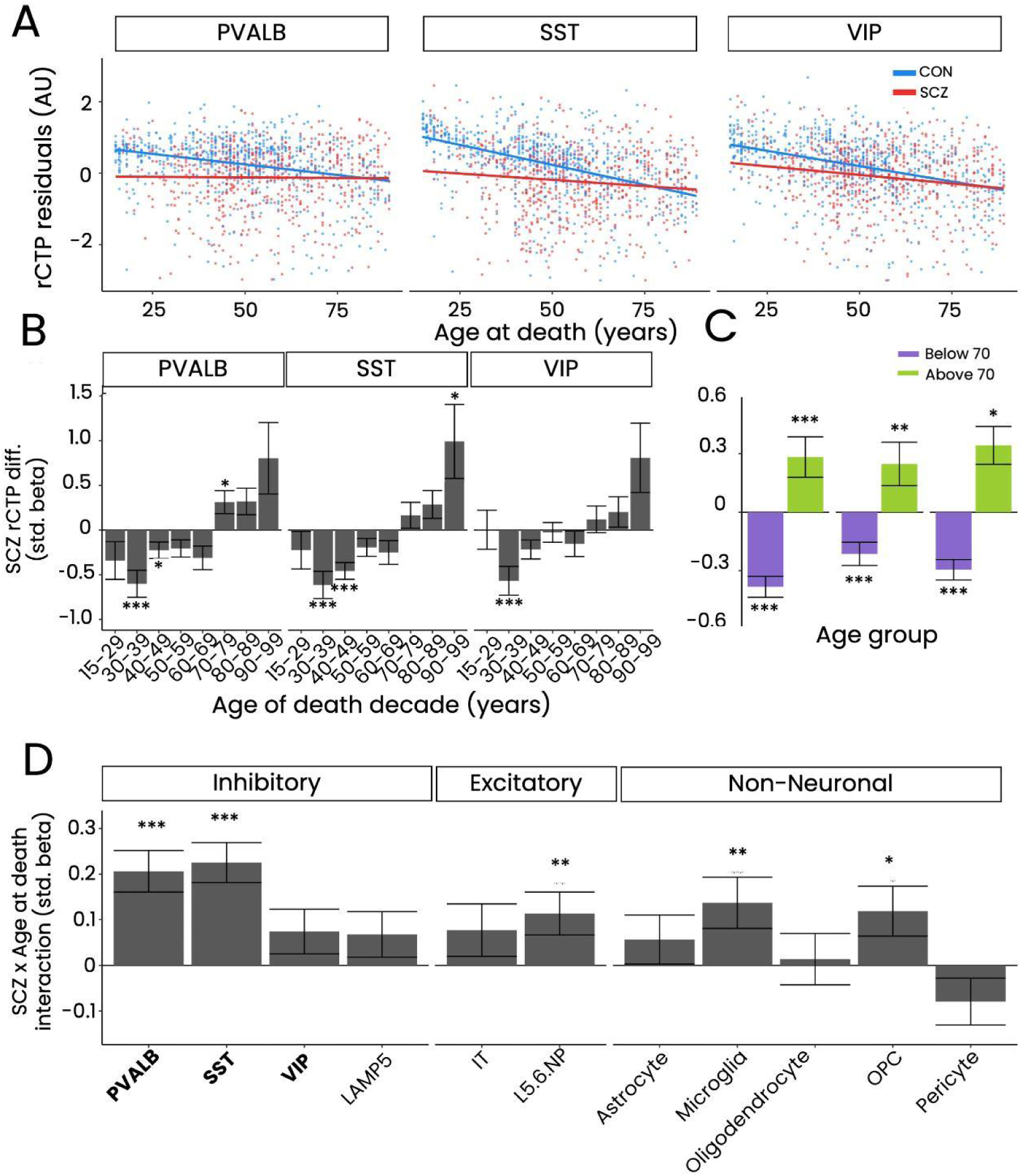
The effect of age at death on schizophrenia-associated differences in DLPFC interneuron proportions. (A) Scatterplots showing the relationship between age at death and residualized proportions of PVALB, SST, and VIP interneurons in schizophrenia (SCZ) and controls. Each point represents an individual bulk RNA-seq sample, aggregated across datasets, with lines indicating the best linear fit for each group. (B) Bar plots displaying the effect of SCZ on interneuron proportions, binned by decade of age at death. The y-axis shows standardized beta coefficients from linear models adjusting for covariates. Error bars represent standard error, and asterisks denote significance based on FDR thresholds (* = 0.1, ** = 0.05, *** = 0.01). (C) Bar plots showing SCZ effects on interneuron proportions for donors above and below 70 years. The y-axis shows beta coefficients from linear models, with error bars and FDR-based significance as in panel (B). (D) Bar plots of the interaction between age at death and diagnosis on cell type proportions. Interaction coefficients indicate how the effect of SCZ on cell type proportions changes with age; positive coefficients show that SCZ-associated differences in cell type proportions increase with age. Error bars represent standard error, and asterisks denote significance based on FDR thresholds.

To quantify age-related differences in GABAergic rCTPs between SCZ cases and controls, we employed two approaches. First, in an age-stratified analysis, we binned donors by decade of death (Figure 3B). We found that SCZ was associated with reduced PVALB, SST, and VIP rCTPs in younger donors, particularly those aged 30-49. This reduction was attenuated in older age bins (50-79), with evidence of increased PVALB and SST rCTPs in the oldest donors (70-99). Similar findings emerged when comparing donors younger than 70 with those older than 70 (Figure 3C), where SCZ cases younger than 70 showed reduced rCTPs, while those older than 70 showed increased rCTPs compared to controls. Second, we tested for an interaction between age at death and SCZ diagnosis, which revealed statistically significant interaction effects for PVALB and SST rCTPs, and weaker interactions for other cell types like L5/6 near-projecting pyramidal cells, OPCs, and microglia.

Second, we tested for an interaction between subject age at death and SCZ case/control diagnosis (see Methods). We observed statistically significant positive interactions between subject age at death and SCZ diagnosis for PVALB and SST rCTPs (Figure 3C). We further observed statistically significant, albeit weaker magnitude, interactions between age and SCZ status for rCTPs for other cell types, including L5/6 near projecting pyramidal cells, microglia, oligodendrocyte precursor cells (OPCs).

In summary, younger donors with SCZ show reduced proportions of PVALB and SST cells, whereas older donors are more likely to exhibit increased proportions compared to controls. These results underscore the critical importance of subject age in determining whether PVALB and SST cell proportions are decreased, unchanged, or even increased in SCZ.

### snRNAseq and FISH-based stereology confirm age-dependent differences in PVALB and SST cell proportions in SCZ

To confirm the SCZ-associated, age-dependent differences in PVALB and SST proportions observed in bulk tissue deconvolution, we analyzed direct measurements of cell type proportions and densities. We used snRNAseq data from DLPFC samples in two cohorts (McLean, MSSM) and FISH-based stereology data from the Pitt cohort, which quantified PVALB, SST, and VIP cell densities in the subgenual ACC (sgACC).

First, snRNAseq data from control donors showed a decrease in snCTPs for older compared to younger donors (Figure 4A), consistent with previous findings. In SCZ cases, those dying before age 70 had reduced PVALB and SST snCTPs compared to controls, while older SCZ donors showed increased snCTPs for these cell types, mirroring our bulk RNAseq results (Figure 4B, C). Importantly, without binning donors by above and below age 70, we see results similar to that of the Ruzicka study (^40^, with no significant associations between SCZ case/control status and interneuron snCTPs (Supplementary Figure S6). As a critical control analysis, none of the other cell types showed a significant interaction between SCZ diagnosis and age at death (Figure 4C). Second, using FISH-based stereology data we had previously collected from the sgACC from donors in the Pitt cohort (Supplementary Figure S7), we confirmed reduced densities in SCZ of PVALB cells (p = 0.025) and statistically trending reductions in SST and VIP cells (p < 0.10).

**Figure 4.**
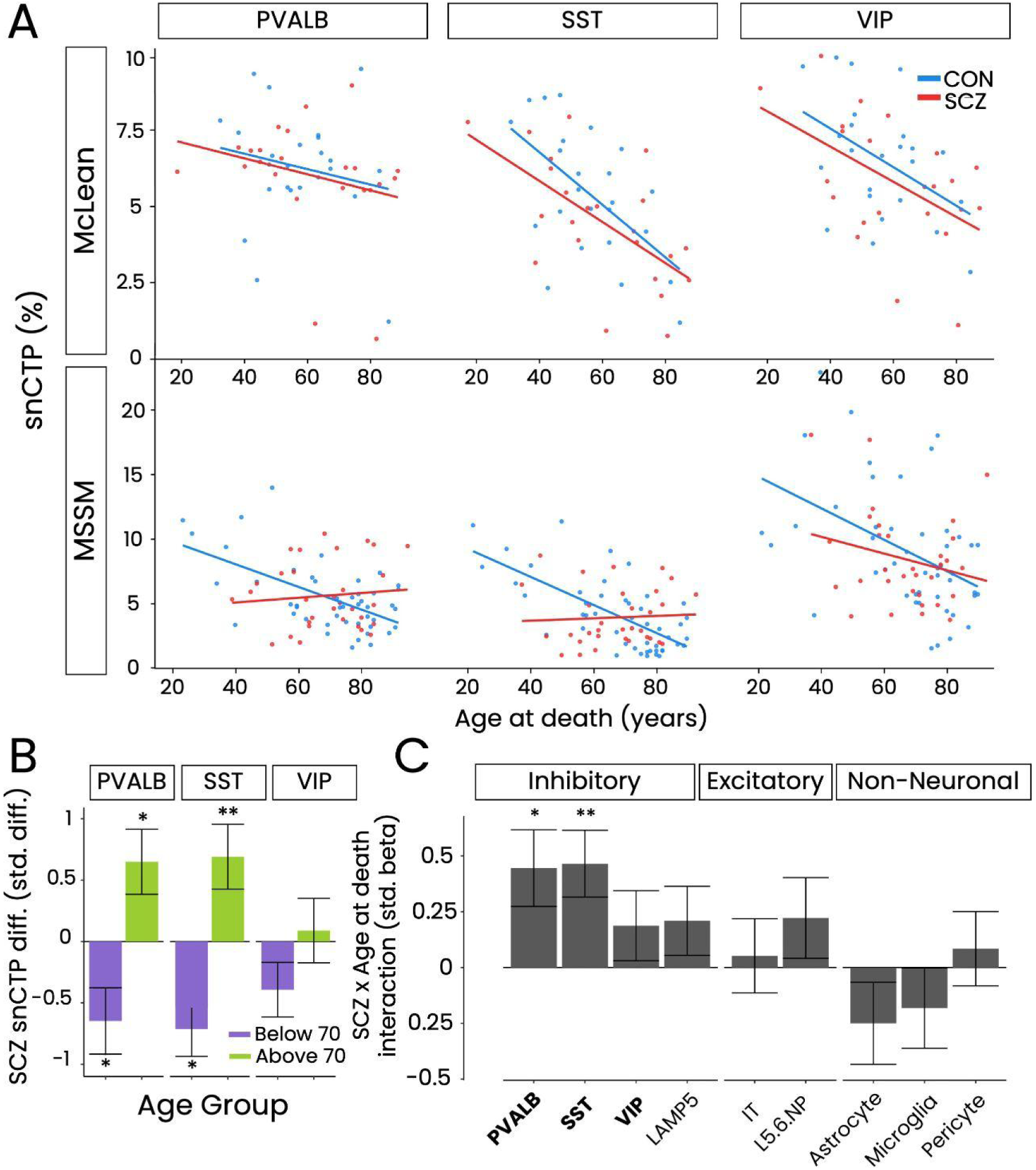
Schizophrenia-associated differences in single-nucleus cell type proportions of DLPFC interneuron subtypes. (A) Scatterplots showing snCTPs for PVALB, SST, and VIP interneurons in schizophrenia (SCZ) and controls, plotted against age at death across two cohorts (McLean and MSSM). Each point represents a snRNAseq sample, with lines showing the best linear fit per diagnosis group. (B) Bar plots of SCZ effects on snCTPs, stratified by age (above or below 70). The y-axis shows standardized beta coefficients from linear models adjusting for covariates. Error bars represent standard error, and asterisks indicate significance based on FDR thresholds. (C) Bar plots showing the interaction between age at death and diagnosis on snCTPs, with significant relationships indicated by bolded cell type labels. Error bars represent standard error, with significance based on FDR thresholds.

In summary, independent analyses using snRNAseq and FISH-based stereology strongly support the age-dependent effects of SCZ on PVALB and SST cell proportions and densities.

### Earlier onset of SCZ is associated with reduced PVALB and SST cell proportions

We next investigated whether clinical factors, such as the age of SCZ onset, were associated with differences in GABAergic interneuron rCTPs. Although detailed clinical or symptom severity information was generally unavailable from donors in these cohorts ^41^, age of SCZ onset was provided in the GVEX and LIBD studies.

After accounting for age at death and other demographic and technical covariates, we found that earlier onset of SCZ was associated with decreased rCTPs in PVALB and SST cells in both the GVEX and LIBD datasets (Figure 5, GVEX: PVALB R = 0.24, p = 0.028; SST R = 0.22, p = 0.047; LIBD: PVALB R = 0.18, p = 0.016; SST R = 0.21, p = 0.0057). Lastly, as SCZ age of onset is expected to be confounded with lifetime duration of SCZ, we note that we did not see significant associations between SCZ duration of illness and PVALB and SST rCTPs (Supplementary Figure S8), suggesting that SCZ age of onset is more related to interneuron rCTPs than lifetime duration of SCZ symptoms.

**Figure 5.**
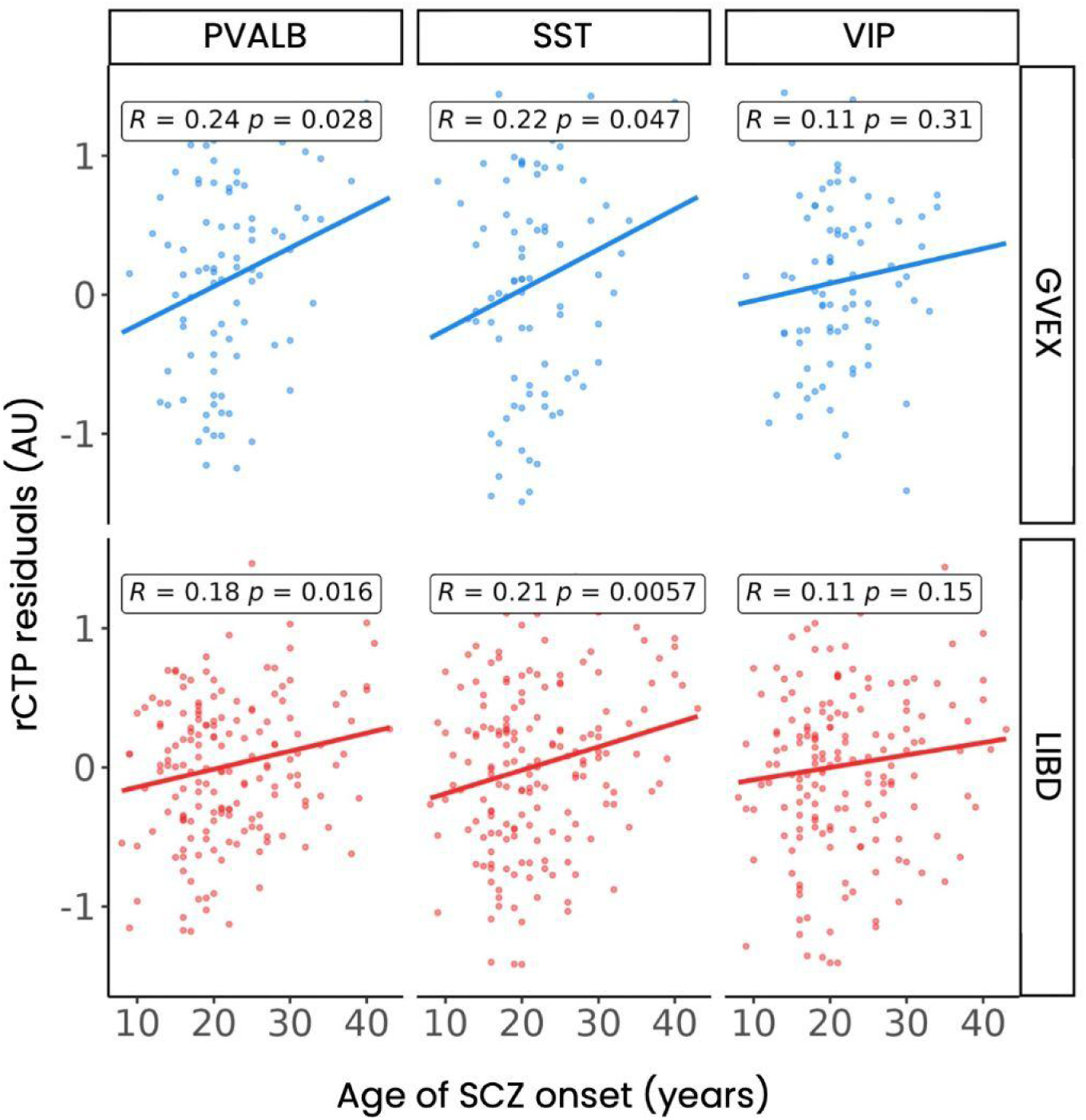
Effects of schizophrenia age of onset on DLPFC interneuron subtype proportions. Associations between age of onset of schizophrenia (SCZ) and residualized cell type proportions (rCTPs) for PVALB, SST, and VIP interneurons. The y-axis shows rCTP estimates for each subtype after residualizing for covariates, including subject age at death. Each point represents a bulk RNA-seq sample, with point color indicating the contributing study (GVEX or LIBD). Inset lines show the best linear fit, with inset Pearson’s R and p-values indicated; p-values not adjusted for multiple comparisons.

These findings show that SCZ cases with an earlier onset of SCZ, and thus potentially more severe symptomatology ^1,10^, are more likely to exhibit greater reductions in PVALB and SST interneurons than donors with later SCZ onset. Hence, among SCZ cases, differences in ages of SCZ onset explains some of the inter-individual heterogeneity in interneuron pathology.

### Cohort-specific transcriptomic dysregulation among GABAergic cell subtypes in SCZ

While our analyses strongly implicate differences in PVALB and SST cell proportions in SCZ, we also explored whether cell type-specific gene expression patterns differ in SCZ. We utilized DLPFC snRNAseq data from the McLean and MSSM cohorts and sgACC LCM-seq data from the Pitt cohort, aiming to replicate findings of Deniel et al. of decreased PVALB and SST mRNA expression in these cells in SCZ ^29^.

We conducted cell type-specific and age-stratified differential gene expression (DEG) analyses, focusing on marker genes that define each major GABAergic cell subtype (see Methods). In the McLean cohort, at a threshold of unadjusted p < 0.05, we found decreased PVALB mRNA expression in PVALB cells in younger SCZ donors (age of death < 70) (Figure 6A, log_2_FC = -0.492, p = 0.044), but no significant differences in older McLean SCZ cases or in either age group in the MSSM cohort. For SST cells (Figure 6B), we observed decreased SST mRNA expression in both younger (log_2_FC = -0.767, p = 0.0425) and older (log_2_FC = -1.85, p = 9.06×10^-3^) donors with SCZ in the McLean cohort, while SST mRNA expression remained unchanged in the MSSM cohort. VIP mRNA expression was generally unchanged (Figure 6C), except for increased expression in older SCZ donors in the MSSM cohort (log_2_FC = 0.732, p = 0.0212).

**Figure 6.**
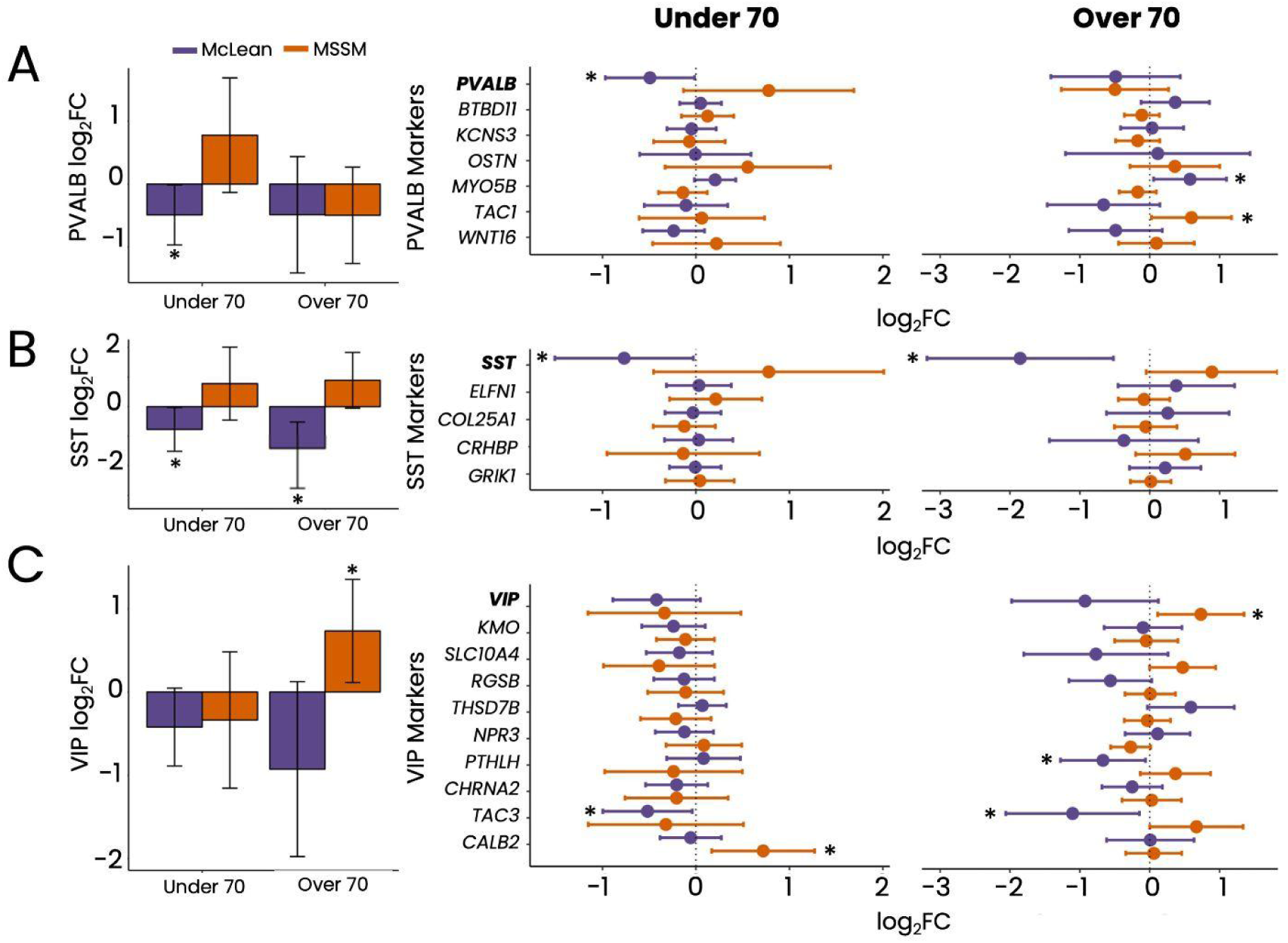
Gene expression differences in DLPFC interneuron marker genes in schizophrenia (SCZ). (A-C) Left panels show SCZ-associated differential expression for PVALB, SST, and VIP mRNA expression in their respective cell types in the McLean and MSSM snRNAseq datasets. Asterisks denote significant differential expression in SCZ (unadjusted p < 0.05). Right panels show differential expression across all cell type-specific marker genes. Bars represent 95% confidence intervals (CI), and asterisks denote significance as in the left panel.

When examining additional marker genes for these cell types (Figure 6A-C, right), we found no consistent patterns of broad up- or down-regulation, suggesting that widespread dysregulation or loss of cell type identity is unlikely in SCZ (see Discussion). In the Pitt cohort’s sgACC LCM-seq data (Supplementary Figure S8), we observed decreased PVALB mRNA expression in PVALB cells in SCZ (log_2_FC = -1.19, p = 0.0196) and reduced but not significant changes in SST (log_2_FC = -0.328, p = 0.141) or VIP (log_2_FC = -0.59, p = 0.065) mRNA expression.

In summary, our findings provide some support for per-cell reductions in PVALB and SST mRNA among PVALB and SST cells in SCZ, though this finding appears somewhat cohort-specific. For example, while we saw evidence for reduced PVALB mRNA among PVALB cells among younger donors in both the McLean and Pitt cohorts, we did not see similar per-cell reductions in PVALB mRNA among younger or older donors in the MSSM cohort. This highlights the need for further investigation of GABAergic cell type-specific dysregulation in additional cohorts with larger sample sizes.

## Discussion

In this study, we leveraged six cohorts encompassing 12 bulk and cell type-specific RNA sequencing datasets to explore the associations between schizophrenia (SCZ) diagnosis and dysregulation of key GABAergic interneuron subtypes in the human DLPFC and ACC. We identified that SCZ was associated with reduced interneuron proportions in younger donors, particularly affecting PVALB and SST cells. However, this relationship attenuated with donor age, and in older donors, particularly those over seventy, we observed a surprising positive association, with increased proportions of these interneurons in SCZ relative to controls. Additionally, we found that an earlier onset of SCZ was associated with greater reductions in PVALB and SST proportions, suggesting that the timing of disease onset played a critical role in determining cellular pathology.

To ensure the robustness of our findings, we employed multiple technologies and methodologies, including bulk RNAseq for cell type deconvolution, fluorescence in situ hybridization (FISH) combined with stereology to validate cellular densities, and snRNAseq and LCM-seq for high-resolution cell-specific insights. This multimodal approach allowed us to cross-validate our results, be less sensitive to outlier cohorts, and gain a more comprehensive understanding of the cellular and molecular changes associated with SCZ.

Our findings are consistent with previous research that has identified disruptions in GABAergic interneurons, particularly PVALB and SST cells, as central to the pathophysiology of SCZ ^26,27,29^. Our study confirms these associations but adds nuance by showing that the direction and magnitude of these changes are highly age-dependent ^33^. While younger SCZ cases exhibit reduced interneuron proportions consistent with prior reports ^26–28^, our observation of increased PVALB and SST proportions in older SCZ patients introduces a new perspective that may reflect compensatory or resilience mechanisms. Moreover, our findings align with the literature on the molecular pathology of SCZ, which has highlighted changes in gene expression within PVALB and SST interneurons ^6,11,28–31^. However, our results also emphasize the variability of these changes across different cohorts, suggesting that cohort-specific factors, such as age distribution or patient-specific symptomatology, may explain some of the discrepancies observed in previous studies.

Among SCZ cases, we found that donors with an earlier age of SCZ onset displayed fewer PVALB and SST cells, suggesting that the developmental stage at which SCZ emerges considerably shapes the degree of interneuron cellular pathology. Early onset SCZ is strongly linked to more severe clinical outcomes, including greater cognitive impairment, more debilitating negative symptoms, and increased hospitalization ^7–10^. Intriguingly, vulnerability of PVALB and SST interneurons may contribute in part to these symptoms, suggesting that early intervention strategies aimed at preserving interneuron health may be particularly beneficial for younger or more severely affected SCZ patients.

One of our most perplexing findings is the observed increase in interneuron proportions in older donors with SCZ relative to controls. This does not suggest that these interneurons are proliferating or ‘growing’ within the brains of these individuals; instead, this likely represents a relative preservation of these cells compared to the typical loss of these cells observed in healthy aging ^22,23^. In other words, older cases with SCZ exhibit less loss of these interneurons than expected when compared to age-matched healthy controls, who would normally exhibit a more pronounced age-related decline in these cells. One explanation for this finding could be potential neuroprotective effects associated with long-term antipsychotic medication use in individuals with SCZ ^55,56^. Alternatively, this finding may reflect a form of survivorship bias, where those donors with SCZ who live into their 70s and beyond may represent an especially healthy subset of the broader SCZ population and thus might be less affected by typical neurodegenerative processes. Moreover, our data indicate that age of onset interacts with age at death, potentially creating distinct disease trajectories for younger-versus older-onset cases, with early onset associated with pronounced interneuron loss and later onset potentially following a slower rate of decline.

Despite the strengths of our study, several limitations warrant acknowledgment. First, our cell type taxonomy, derived from snRNAseq data of healthy donors ^43,46^, may not fully represent the cellular landscape of SCZ or account for age-related changes in disease states. Additionally, our use of bulk RNAseq deconvolution provides a useful but indirect proxy for cell proportions ^57^, despite our careful benchmarking against “gold-standard” snRNAseq cell counts and stereology-derived cell densities. Another challenge is the cohort-specific nature of some of our findings; variations in factors such as subject age at death and subtle differences in diagnostic criteria among cohorts may affect the observed associations. Moreover, while our study illuminates the age-dependent effects of SCZ on interneurons, it does not fully explore the potential roles of other cell types, like pyramidal cells, which are also critical to SCZ pathology ^58^. Furthermore, our reliance on postmortem data limits our ability to infer dynamic pathological processes occurring within individuals over their lifetimes.

Our study has several important implications for future research. The age-dependent changes in interneuron proportions suggest that age-specific interventions could be beneficial in treating SCZ. Future studies should explore whether targeted therapies aimed at preserving or enhancing PVALB and SST function are more effective in younger or older SCZ patients. Additionally, the potential neural resilience observed in older SCZ donors warrants further investigation. Identifying the factors that contribute to this resilience could lead to new therapeutic strategies aimed at mitigating the severity of SCZ. The observed variability across cohorts highlights the need for ongoing large-scale, diverse studies, such as PsychENCODE ^41,59^, that can account for demographic and clinical differences. Furthermore, given the potential impact of interneuron changes on cortical microcircuit function, future studies employing computational models ^60,61^, could investigate how cellular alterations in these specific cell types affect information processing and cognitive functions in SCZ patients. Better understanding these mechanisms could provide deeper insights into the cognitive symptoms of SCZ and inform the development of interventions aimed at restoring normal cortical function.

In conclusion, our study provides new insights into the complex, age-dependent nature of interneuron dysregulation in SCZ and underscores the need for a multifaceted approach to understanding and treating this disorder.

## Data Availability

All data produced in the present study are available upon reasonable request to the authors and can also be requested through the PsychENCODE consortium.

https://www.synapse.org/#!Synapse:syn4921369

## Acknowledgements

This work was supported generous support from the CAMH Discovery Fund, Krembil Foundation, Natural Sciences and Engineering Research Council of Canada (RGPIN-2020-05834 and DGECR-2020-00048), Canadian Institutes of Health Research (PJT-191747, NGN-171423, and PJT-175254), the Simons Foundation Autism Research Initiative, and Brain Canada. We are also grateful for donors and donors’ families for donation of brain tissues without which this work would not be possible. We acknowledge useful discussions and feedback from Paul Pavlidis, Lilah Toker, and the Tripathy and Sibille labs and members of the Krembil Centre for Neuroinformatics.

We acknowledge the use of datasets from the CommonMind and PsychEncode Consortia. The CommonMind Consortium was supported by funding from Takeda Pharmaceuticals Company Limited, F. Hoffman-La Roche Ltd and NIH grants R01MH085542, R01MH093725, P50MH066392, P50MH080405, R01MH097276, RO1-MH-075916, P50M096891, P50MH084053S1, R37MH057881, AG02219, AG05138, MH06692, R01MH110921, R01MH109677, R01MH109897, U01MH103392, and contract HHSN271201300031C through IRP NIMH. Data were generated as part of the PsychENCODE Consortium, supported by: U01DA048279, U01MH103339, U01MH103340, U01MH103346, U01MH103365, U01MH103392, U01MH116438, U01MH116441, U01MH116442, U01MH116488, U01MH116489, U01MH116492, U01MH122590, U01MH122591, U01MH122592, U01MH122849, U01MH122678, U01MH122681, U01MH116487, U01MH122509, R01MH094714, R01MH105472, R01MH105898, R01MH109677, R01MH109715, R01MH110905, R01MH110920, R01MH110921, R01MH110926, R01MH110927, R01MH110928, R01MH111721, R01MH117291, R01MH117292, R01MH117293, R21MH102791, R21MH103877, R21MH105853, R21MH105881, R21MH109956, R56MH114899, R56MH114901, R56MH114911, R01MH125516, and P50MH106934 awarded to: Alexej Abyzov, Nadav Ahituv, Schahram Akbarian, Alexander Arguello, Lora Bingaman, Kristin Brennand, Andrew Chess, Gregory Cooper, Gregory Crawford, Stella Dracheva, Peggy Farnham, Mark Gerstein, Daniel Geschwind, Fernando Goes, Vahram Haroutunian, Thomas M. Hyde, Andrew Jaffe, Peng Jin, Manolis Kellis, Joel Kleinman, James A. Knowles, Arnold Kriegstein, Chunyu Liu, Keri Martinowich, Eran Mukamel, Richard Myers, Charles Nemeroff, Mette Peters, Dalila Pinto, Katherine Pollard, Kerry Ressler, Panos Roussos, Stephan Sanders, Nenad Sestan, Pamela Sklar, Nick Sokol, Matthew State, Jason Stein, Patrick Sullivan, Flora Vaccarino, Stephen Warren, Daniel Weinberger, Sherman Weissman, Zhiping Weng, Kevin White, A. Jeremy Willsey, Hyejung Won, and Peter Zandi.

## Supplementary Figures

**Figure S1.**
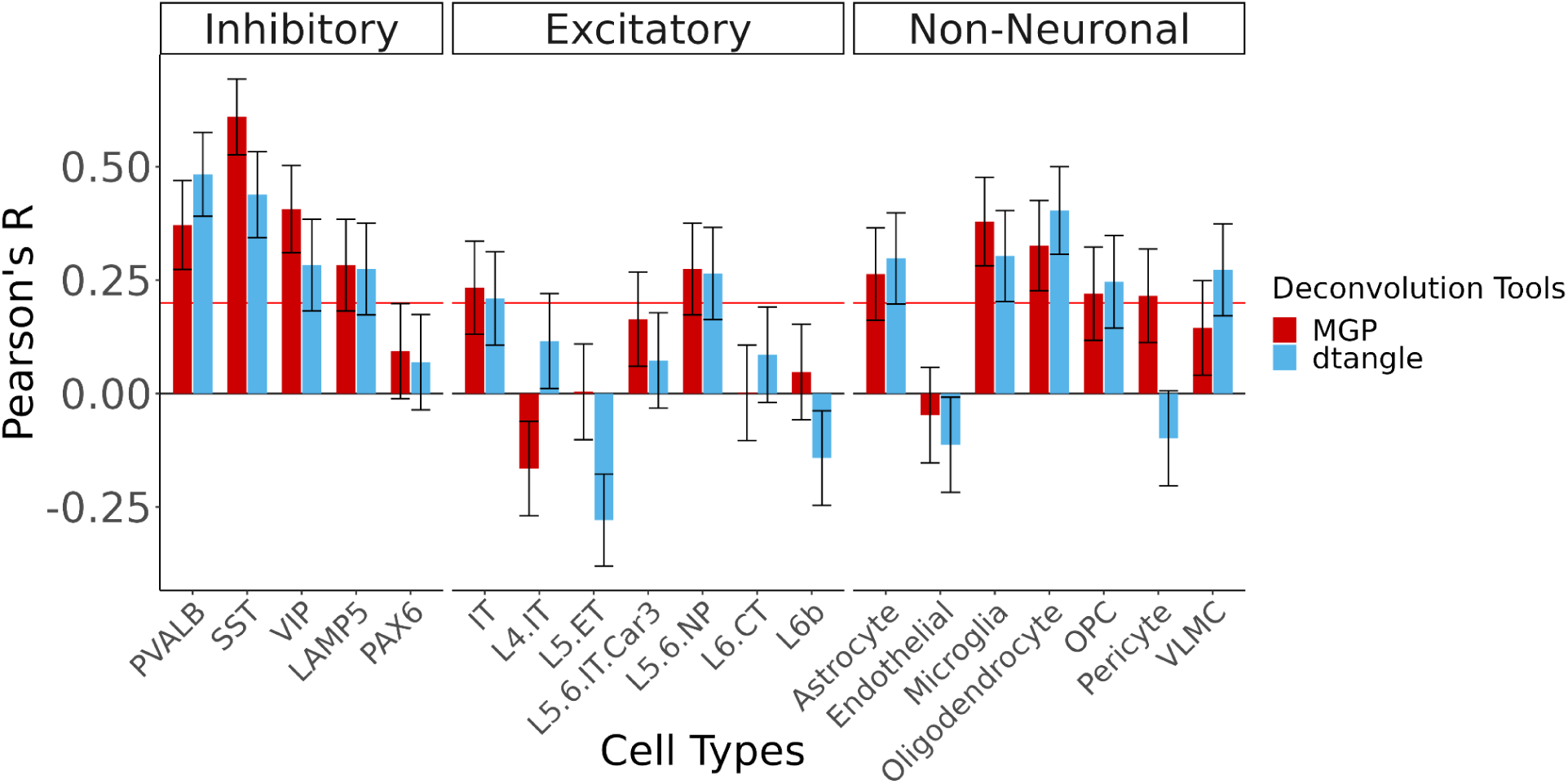
Assessment of Cell Type Proportion Predictions from dtangle against snCTPs from matched donors. Barplot depicting Pearson’s correlations (R) between dtangle-derived bulk deconvolution estimates and single-nucleus cell type proportions. Higher values indicate better concordance between bulk deconvolution and single-nucleus cell type proportions. Error bars represent the standard error for each Pearson correlation.

**Figure S2.**
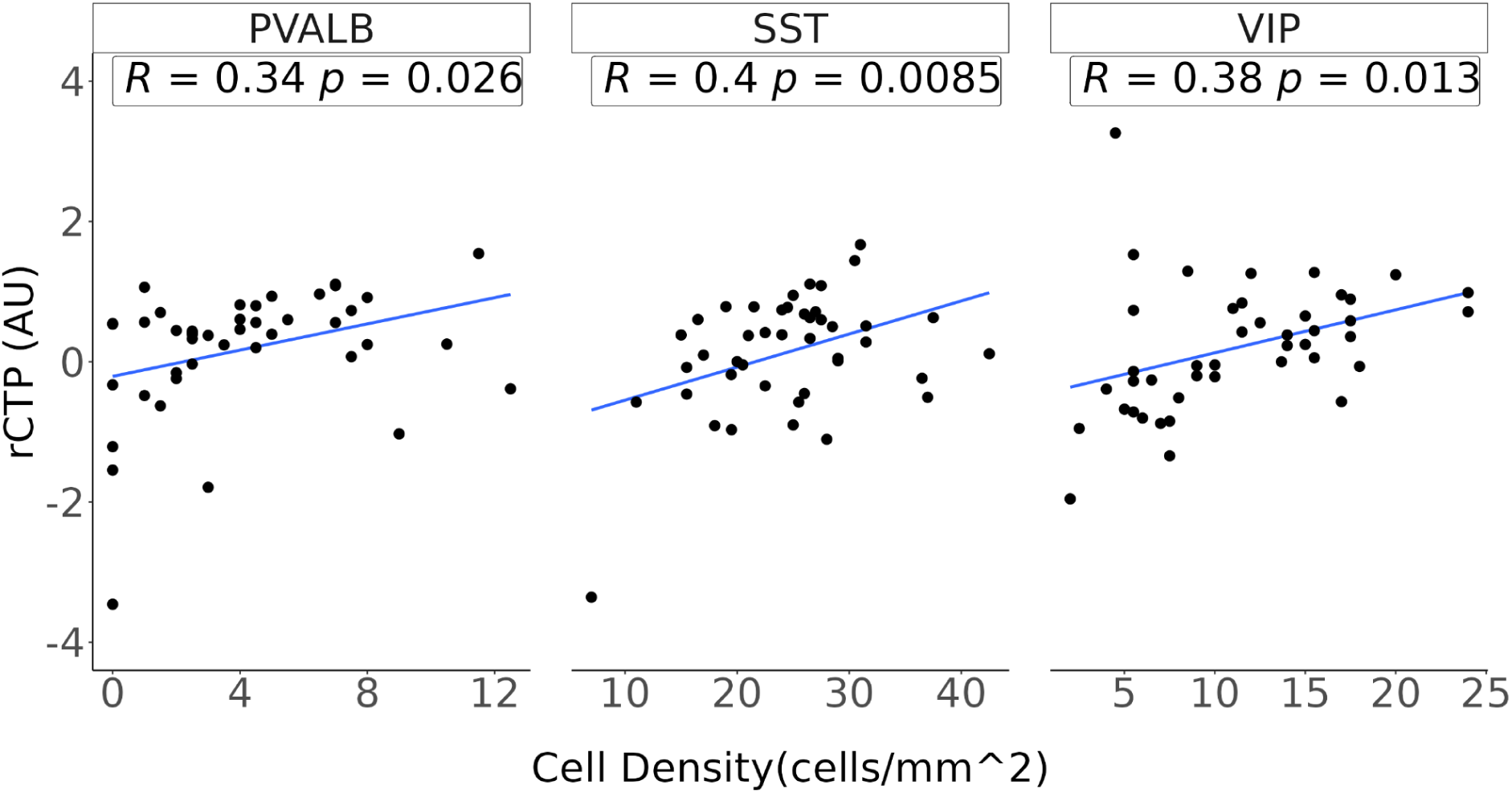
Correlation of MGP-derived rCTPs with microscopy-based cell densities for three major interneuron types in ACC from matched donors. (B) Scatterplots showing the correlation between fluorescence in situ hybridization-based cell densities (expressed as number of labelled cells per millimeter squared, x-axis) and relative cell type proportions (rCTPs; y-axis, arbitrary units, AU) derived from bulk tissue deconvolution via the MGP algorithm. Inset values show Pearson’s correlation coefficient (R) and corresponding p-values.

**Figure S3.**
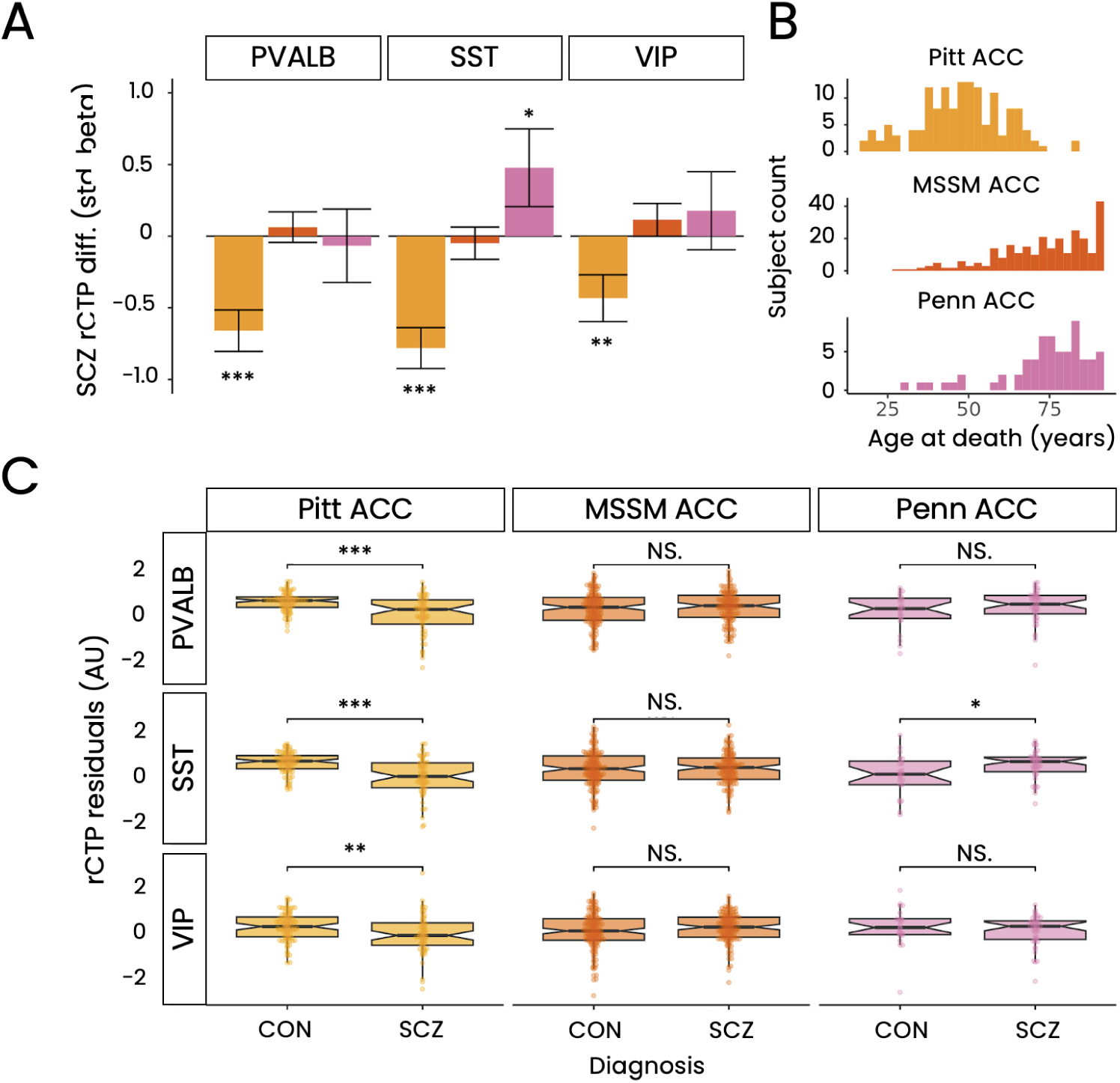
Differences in ACC interneuron proportions between schizophrenia and controls. (A) Bar plots showing standardized beta coefficients (β) for differences in relative cell type proportions (rCTPs) of PVALB, SST, and VIP interneurons between schizophrenia (SCZ) and control (C) groups across three bulk ACC RNA-seq datasets. Positive values indicate increased proportions in SCZ, while negative values indicate decreases. Error bars represent standard error, and asterisks indicate significance based on false discovery rate FDR (* = 0.1, ** = 0.05, *** = 0.01). (B) Age at death distribution for each dataset. Datasets are ordered by increasing mean age in years. (C) Box plots of residualized rCTPs, after controlling for demographic and technical covariates, for PVALB, SST, and VIP interneurons across datasets. Asterisks reflect significance from panel (A), with “NS” for non-significant comparisons.

**Figure S4.**
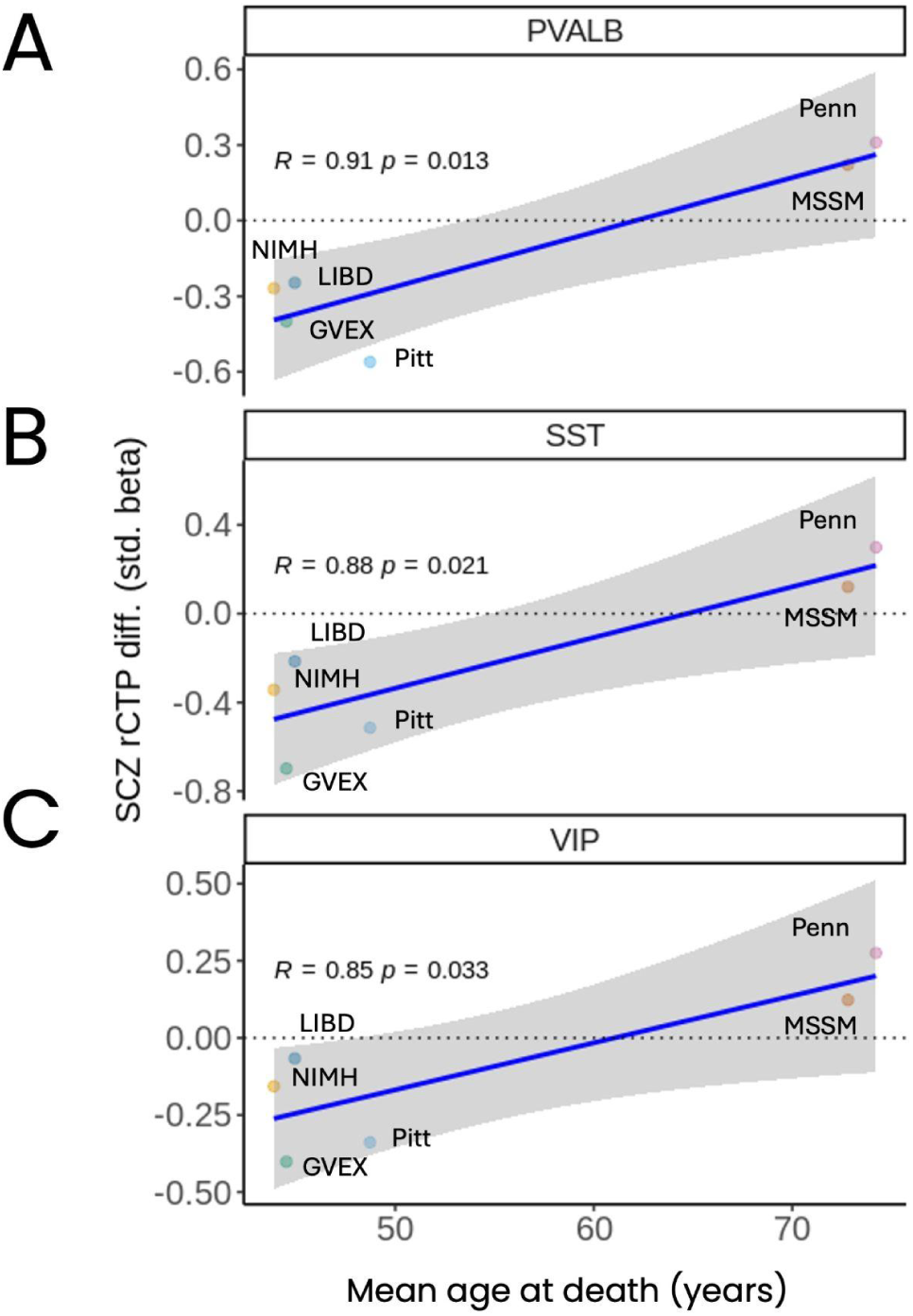
Correlating average DLPFC dataset age at death with SCZ-rCTP association strength. (A-C) Associations between the degree to which SCZ is associated with reduced or increased cell type proportions of PVALB, SST, and VIP cells and mean dataset age at death in size PsychEncode DLPFC datasets. Y axis shows standard beta coefficient values (as in Figure 2A) based on linear modelling and accounting for covariates. X-axis shows the mean age at death for each cohort. Cohorts are marked and labelled on each plot. Inset line represents linear model fit, and shaded area indicates standard error.

**Figure S5.**
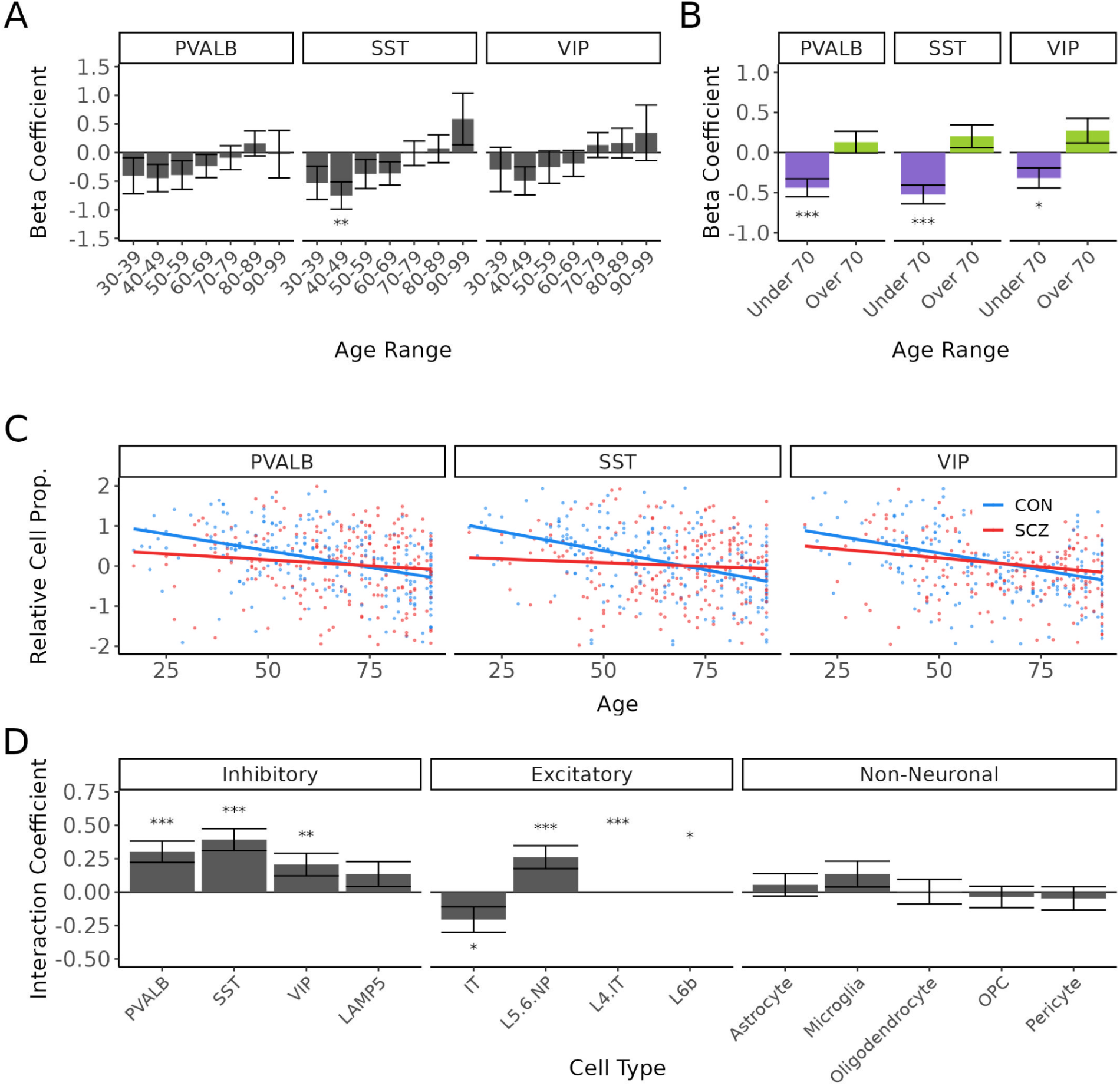
The effect of age at death on schizophrenia-associated differences in ACC interneuron proportions. (A) Scatterplots showing the relationship between age at death and residualized proportions of PVALB, SST, and VIP interneurons in schizophrenia (SCZ) and controls. Each point represents an individual bulk RNA-seq sample, aggregated across ACC datasets, with lines indicating the best linear fit for each group. (B) Bar plots displaying the effect of SCZ on interneuron proportions, binned by decade of age at death. The y-axis shows standardized beta coefficients from linear models adjusting for covariates. Error bars represent standard error, and asterisks denote significance based on FDR thresholds (* = 0.1, ** = 0.05, *** = 0.01). (C) Bar plots showing SCZ effects on interneuron proportions for donors above and below 70 years. The y-axis shows beta coefficients from linear models, with error bars and FDR-based significance as in panel (B). (D) Bar plots of the interaction between age at death and diagnosis on cell type proportions. Interaction coefficients indicate how the effect of SCZ on cell type proportions changes with age; positive coefficients show that SCZ-associated differences in cell type proportions increase with age. Error bars represent standard error, and asterisks denote significance based on FDR thresholds.

**Figure S6.**
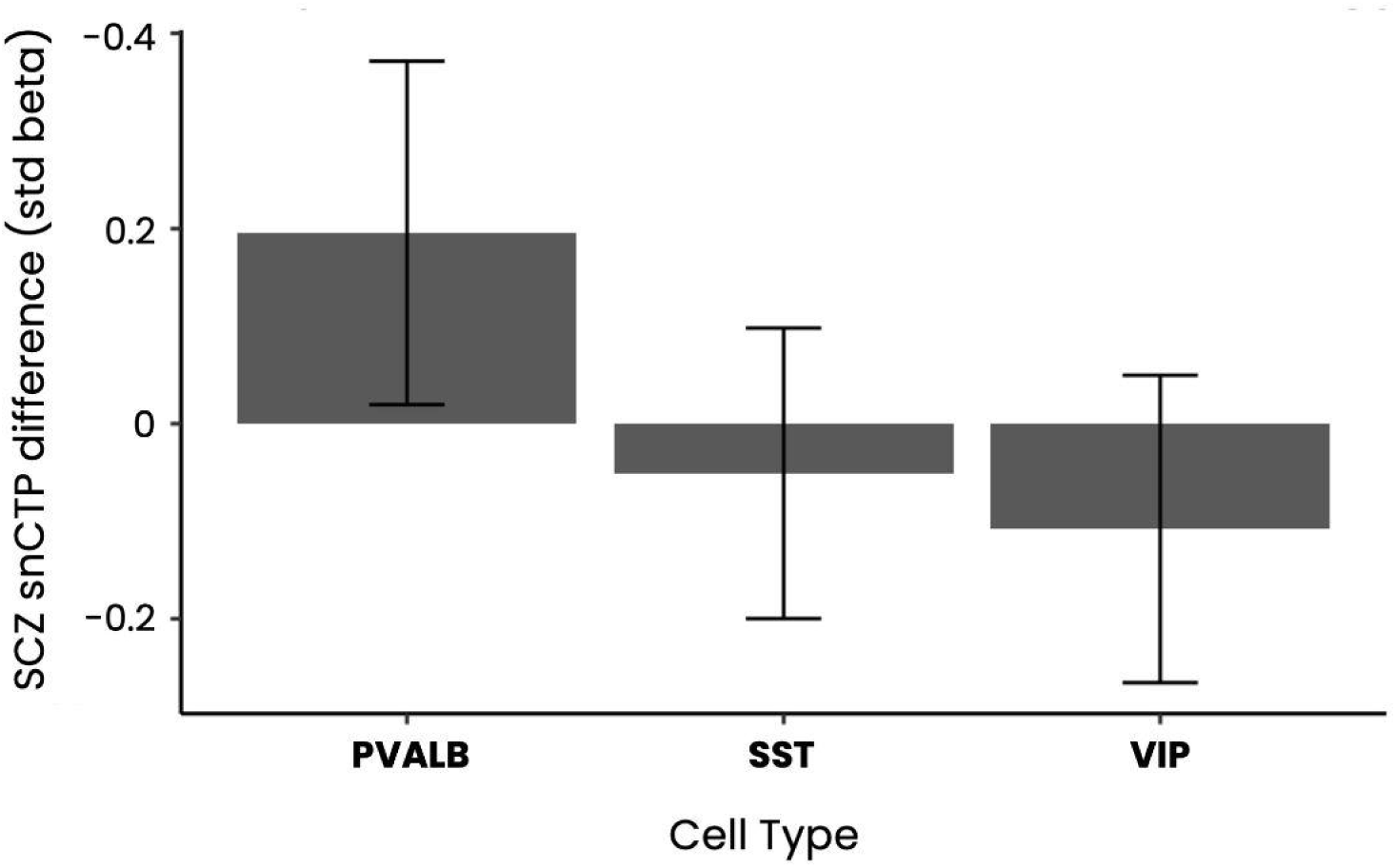
Schizophrenia-associated differences in single-nucleus cell type proportions of DLPFC interneuron subtypes without stratified age groups. Bar plots of SCZ effects on snCTPs without stratifying for age. The y-axis shows standardized beta coefficients from linear models adjusting for covariates. Error bars represent standard error, and asterisks indicate significance based on FDR thresholds, with no asterisk indicating FDR > 0.01.

**Figure S7.**
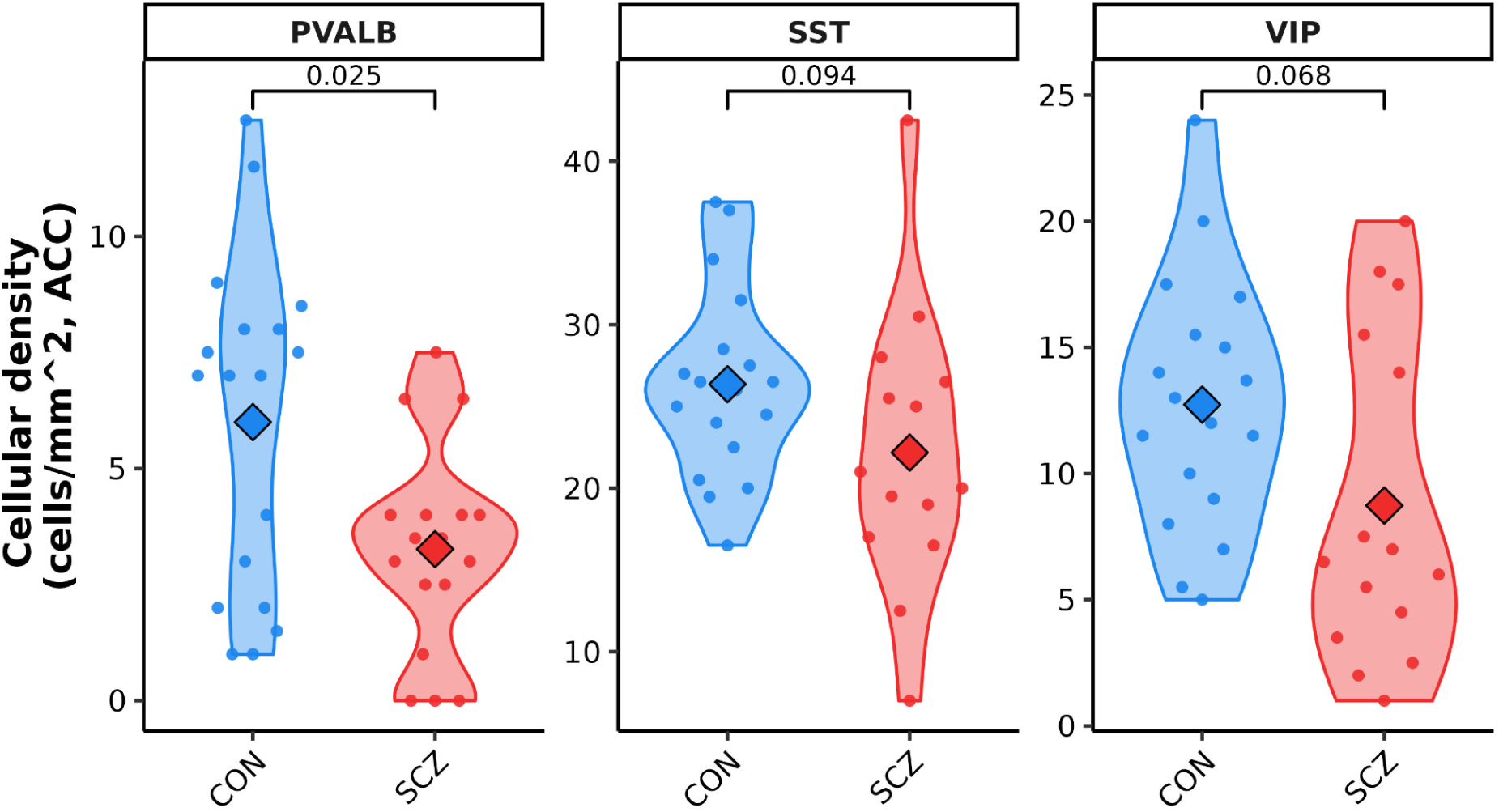
Differences in fluorescence in situ hybridization-based stereological densities of sgACC interneuron subtypes in control and schizophrenia samples. Panels show densities of PVALB, SST, and VIP interneurons stained using double-label fluorescent in situ hybridization (FISH) on sgACC sections. Dots reflect per-subject quantification of counts of labelled cells expressing more than 10 or more cell type-specific marker grains per unit area. Outlier samples removed independently per group if outside range defined by 3 times the interquartile range. Inset p-values denote Wilcoxon signed-rank test.

**Figure S8.**
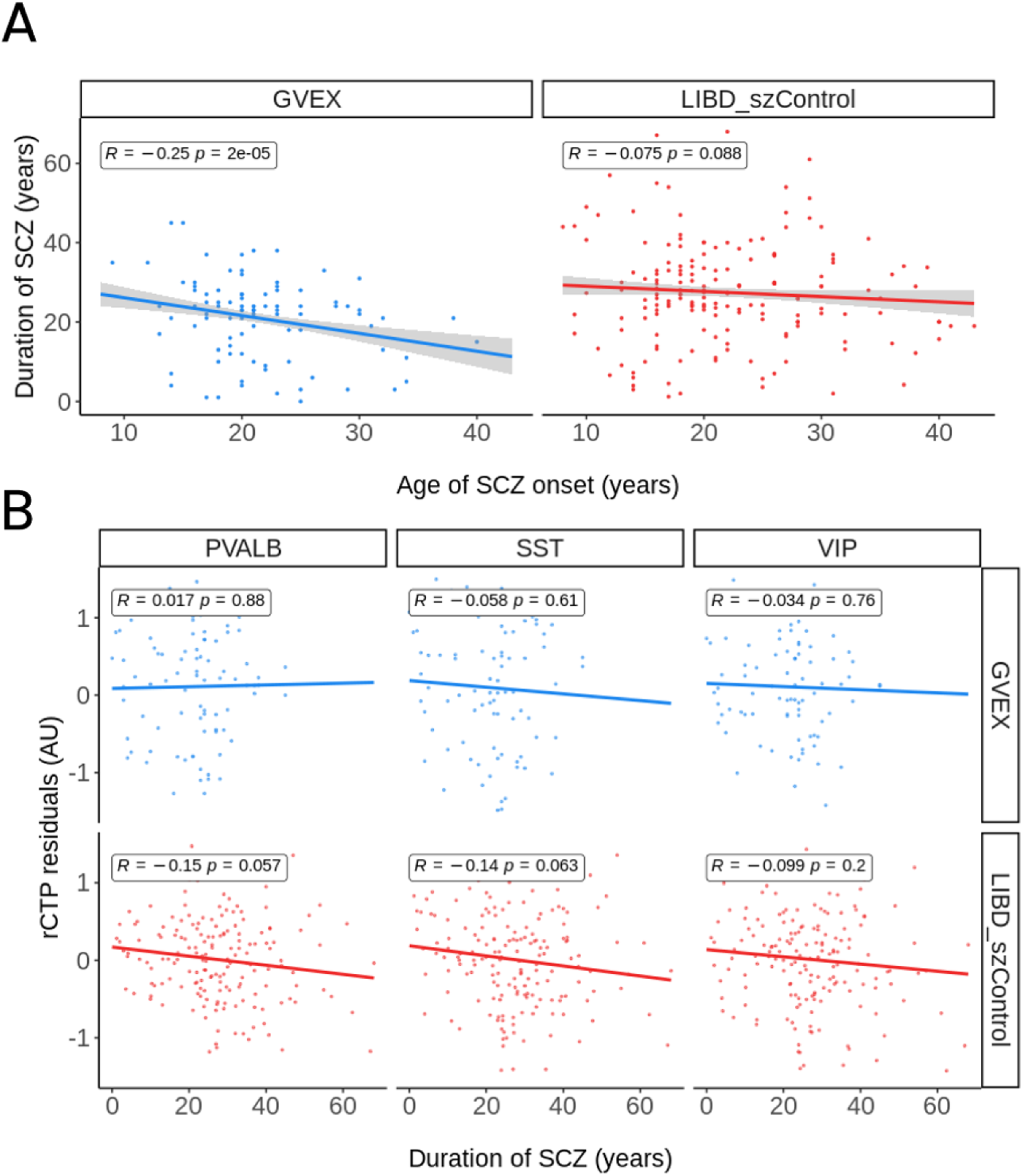
Associations between duration and age of schizophrenia onset with interneuron cell type proportions. (A) Associations between SCZ duration of illness, quantified as years since SCZ age of onset and age at death, and age of SCZ onset in each dataset where SCZ age of onset information was available. Dots reflect distinct donors and Y-axis shows SCZ duration of illness, and X-axis shows age of SCZ onset for each individual in each dataset. Inset line shows best linear fit, shading shows standard error, and corresponding Pearson’s R and p-values are indicated. (B) Associations between SCZ duration of illness and relative cell type proportions for interneuron subtypes in the GVEX and LIBD datasets. Y-axis shows rCTP estimates for SST, PVALB, and VIP interneurons, after residualizing for age of death and other covariates. Each point indicates one post-mortem bulk RNAseq sample and point color indicates contributing study (GVEX or LIBD_szControl). Inset line shows best linear fit and corresponding R and p-values are indicated.

**Figure S.**
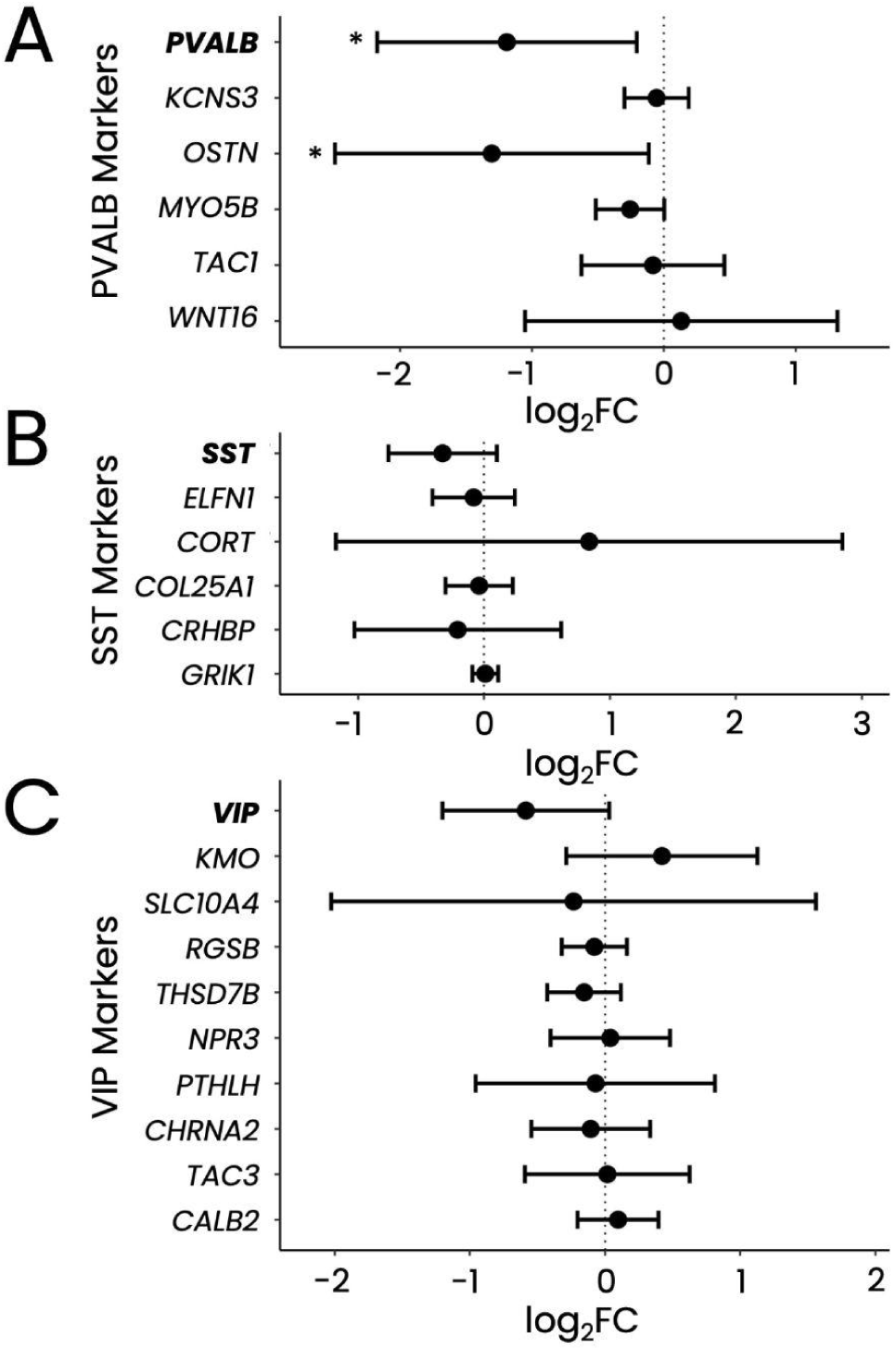
LCM-seq based gene expression differences in sgACC interneuron marker genes in schizophrenia (SCZ). (A-C) SCZ-associated differential expression for PVALB, SST, and VIP interneurons (A-C respectively) in the Pitt LCM-seq dataset. Each panel shows differential expression across all cell type-specific marker genes. Bars represent 95% confidence intervals (CI), and asterisks denote significance (unadjusted p<0.05).

## Notes

### Competing Interest Statement

The authors have declared no competing interest.

### Funding Statement

This study did not receive any funding.

### Author Declarations

The study used data available through the PsychENCODE consortium located at https://www.psychencode.org/

